# RObotic WAlking for children who CAnnot WAlk (RoWaCaWa): Impacts on Physical Function and Physical Activity from a 12-week robotic walking intervention

**DOI:** 10.64898/2026.08.24.26361255

**Authors:** Jessica L. Youngblood, Christa M. Diot, Benjamin M. Norman, Karin Eldred, Amanda Rande, Sean P. Dukelow, Hana Alazem, Anna McCormick, Patricia E. Longmuir, Hua Shen, Kelly A. Larkin-Kaiser, Elizabeth G. Condliffe

## Abstract

**Purpose:** To explore how 12-weeks of robotic walking impacts physical function and sequelae of inactivity for individuals with pediatric-onset neuromotor impairments.

**Methods:** A single-arm mixed-methods interventional study examined robotic walking for 12-weeks in home and community settings, with 12-week follow-up. Outcomes included family goals (Goal Attainment Scale (GAS)) and perspectives (Interviews), postural control (Early Clinical Assessment of Balance), physical activity (Actigraphy, Habitual Activity Estimation Scale, Patient Reported Outcome Measurement Information System (PROMIS) Physical Activity) and sequelae of inactivity (PROMIS Sleep Disturbances, Bowel Function Diary). GAS was collected pre-training, post-training, and 12-week follow-up. All other quantitative outcomes were collected every 4-weeks. Quantitative data are described with median (25^th^-75^th^percentile) and analyzed using a Skillings-Mack test with post-hoc Wilcoxon Signed-Rank. Qualitative interviews were conducted before and after training and analyzed thematically.

**Results:** 15 participants aged 4-23 completed this study. Participants had cerebral palsy (10/15) or rare genetic conditions (5/15), and most used a wheelchair in community settings. Postural control improved (test-statistic = 23.0, p<0.001) after 8 weeks (change=5.0(0.0-21.4), p=0.016) and was maintained through 12-week follow-up (change=13.7(3.1 – 23.7), p=0.008). Over half of the participants achieved goals (t-score > 50) after training. Exploratory analyses suggest improvements in sleep disturbance immediately after training (p=0.025) and 4-weeks after (p=0.047). All measures of physical activity did not improve. Parents reported improvements in walking, activities of daily living, and sequelae of inactivity (i.e., bowel function, appetite, and sleep).

**Conclusions:** Improvements were seen across a range of measures and notably postural control improvements were maintained at the follow-up. Parents perceived improvements in physical function and activities of daily living. Future research is warranted to further understand the impacts of robotic walking for children and small adults with mobility impairments.

## Introduction

Many children (∼1/1000) are unable to walk due to cerebral palsy (Amankwah et al., 2020; Smithers-Sheedy et al., 2016) or other neuromotor impairments (e.g. genetic condition, spina bifida, and acquired brain injuries). These individuals also experience muscle weakness, impaired motor control, and difficulties with coordination and postural control that greatly impact their daily lives (Bekteshi et al., 2023; Clewes et al., 2024). Those unable to walk spend the majority of their days sedentary (Martin Ginis et al., 2021). Living a sedentary lifestyle can lead to important metabolic consequences and ultimately chronic health conditions (Verschuren et al., 2014). Unfortunately, there are few interventions designed to increase mobility, improve motor function or increase physical activity in children with the most severe neuromotor impairments. Robotic walking is an exciting new technology that has the promise to improve many aspects of physical function for individuals with neuromotor impairments.

Current therapy recommendations for children living with neuromotor impairments include increasing gait training and physical activity during rehabilitation sessions (Demont et al., 2022). However, the gait and strength training done in rehabilitation settings has mostly been effective among those with less severe mobility impairments (GMFCS I – III) (Bekteshi et al., 2023). Body weight-assisted treadmill training and some robotic walkers (e.g. Lokomat) have been used to increase walking in children with severe mobility impairments (Mattern-Baxter, 2009). However, using this technology in a clinical setting can increase workload for clinicians and cause strain on the health care system (Cao et al., 2021; Swank et al., 2020). Beyond walking, there are many other important aspects to address in rehabilitation sessions, such as improving postural control and sequelae of inactivity (e.g. bowel function and sleep). There are limited therapies to address these aspects, and many that do exist are typically only done in children GMFCS I-III (Noritz et al., 2022; Novak et al., 2020). Postural control is important for activities of daily living, but it is impaired in individuals with neuromotor impairments, and more interventions are needed to improve this (Toohey et al., 2024). Robotic walking may address many of the limitations of current rehabilitation programs, and implementing robotic walking devices in a real-world setting may allow for more intense and frequent rehabilitation, as it eliminates the need for staff to be available, and families do not need to travel to appointments.

Robotic walking provides reciprocal stepping in a set gait pattern that can significantly improve motor function (Borggraefe et al., 2010). Robotic walking is advantageous because individuals can receive a high dose of therapy with reduced staffing and resources compared to a clinical setting (Pool et al., 2021). Robotic walking has been shown to have a variety of benefits, such as improvements in gross motor function, walking speed, participant goals, and a significant decrease in sedentary time (Kim et al., 2021; Livingstone & Paleg, 2016; Wright et al., 2021). These improvements are not consistent in all studies, and more work is needed to understand a wider variety of impacts these devices can have (van Hedel et al., 2016). The Trexo robotic walker (Trexo Robotics, Mississauga, ON, CA) is designed to be used by children and small adults with severe mobility impairments and has been shown to improve goals created by parents (measured using the Goal Attainment Scale (GAS)), sleep, and physical activity (Bradley et al., 2026; Diot et al., 2023; Hilderley et al., 2026; Youngblood et al., 2025). Further, it is also important to note that children report improved enjoyment when using the Trexo in comparison with other therapies(Bradley et al., 2026; Youngblood et al., 2026). Most current studies examining robotic walking have focused on motor function and gait outcomes. However, there is still little known about the wide variety of impacts robotic walking may have on physical function, physical activity and what improvements parents notice from using these devices.

The purpose of this study was to understand how 12-weeks of robotic walking would impact the physical function and inactivity sequelae experienced by individuals living with mobility impairments. This was an exploratory study; however, our hypothesis was that individuals doing regular robotic walking would improve in parent-directed goals and improve in multiple aspects related to physical function and physical activity.

## Methods

### Study Design

**Ro**botic **Wa**lking for Children Who **Ca**nnot **Wa**lk (RoWaCaWa) was a single-arm interventional study conducted to understand how robotic walking in home and community settings for 12 weeks impacts family goals, physical function, and sequelae of inactivity. We used a triangulation mixed-methods approach to examine the quantitative and qualitative impacts simultaneously. Evaluating the feasibility of the study methods was one of the objectives (previously reported) (Youngblood et al., 2026). Participants were asked to commit to the intervention and to monthly in-person assessments. All other outcomes were exploratory, and participants could opt out. Ethics approval was obtained from the University of Calgary Conjoint Health and Research Ethics Board (REB21-1166), and written informed consent was obtained from all participants’ legal guardians. The trial was registered with ClinicalTrials.gov (ID: NCT05473676). This study included many outcomes determined through a pragmatic approach; in order to reduce the burden on parents, participants had the option not to participate in outcomes they felt were not feasible for them. The study protocol and outcomes related to feasibility and family impacts have been reported previously (Youngblood et al., 2026).

### Device Description

We used a Trexo robotic walker (Mississauga, ON, CA) in this study, which consists of two robotic legs with motors aligned bilaterally at the hips and knees, attached to a Rifton Pacer (Community Product LLC, Rifton, NY, US) (Figure 1). This device is designed for users who weigh < 150 lbs and have femur lengths of < 35 cm and calf lengths of < 43 cm. The devices were configured in multiple ways to individualize them to fit the needs of each participant. This device is connected to a tablet, which is used to control the cadence, gait mode, joint forces, and range of motion. A total of six robotic walkers were used in this study (3 large and 3 medium).

**Figure 1:**
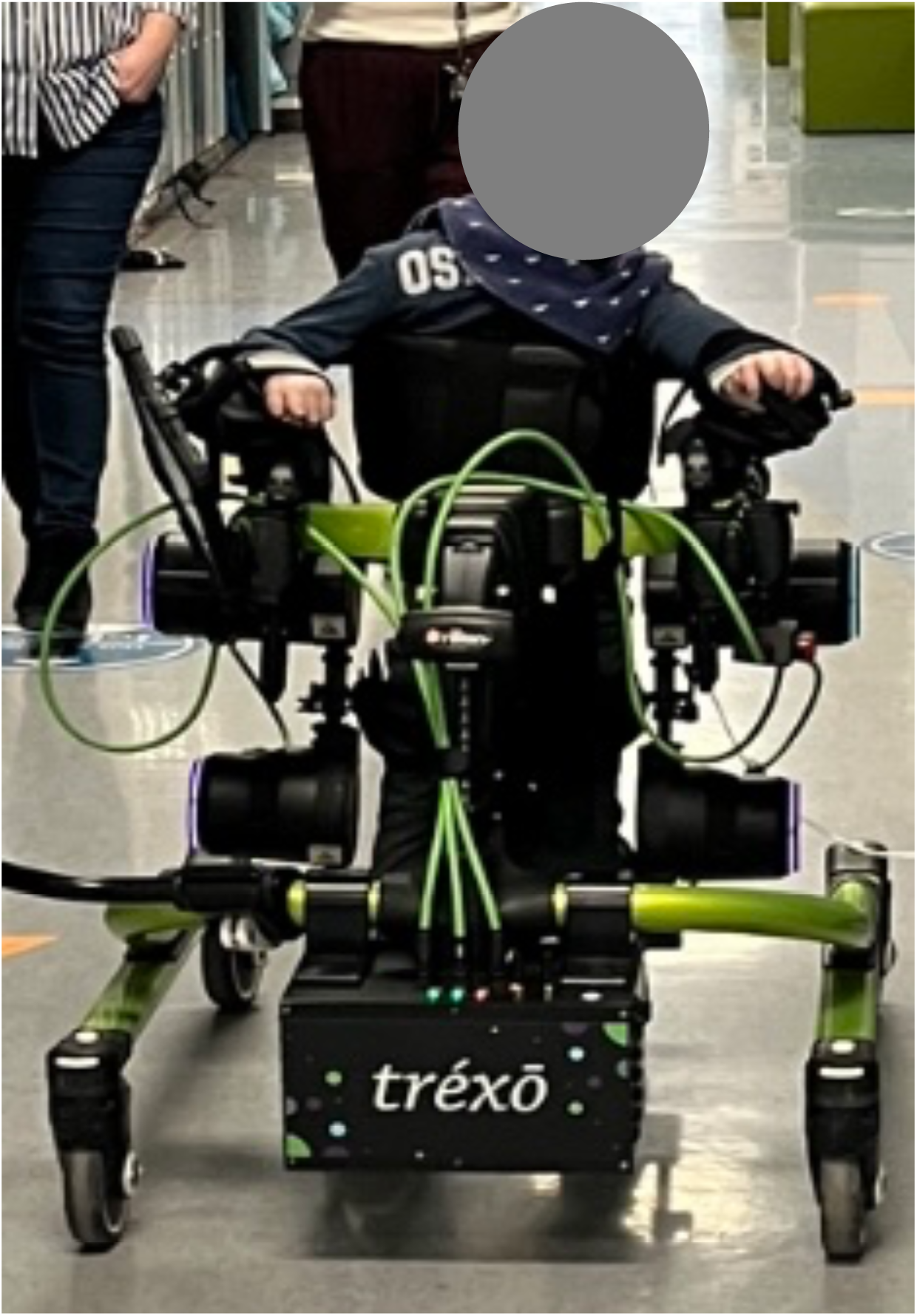
Participant facing anteriorly in a Trexo. This photo is shared with permission.

### Participants

Potential participants self-identified after hearing about the study through community members or social media or were referred by treating clinicians. Before being enrolled in the study, all participants completed a trial with the robotic walker to ensure they met the inclusion criteria and fit the sizes of robotic walkers available. Inclusion criteria were: unable to walk independently in community settings (i.e. Functional Mobility Scale at 500 m of N, C or 1) due to pediatric-onset, non-progressive central nervous system disorder or injury (e.g. cerebral palsy, genetic conditions, spina bifida), at least four years of age, able to fit in a Trexo, and able to comply with study procedures (assessments and training). Exclusion criteria were: participants had a medical condition or recent surgery requiring physical activity restriction (e.g. unstable arrhythmia) or lower extremity immobilization or weight-bearing restrictions (e.g. fracture, unstable hip subluxation), pain or symptomatic hypotension while standing, contracture such that the Trexo robotic gait trainer did not result in forward movement, or simultaneous involvement in a potentially confounding intervention. Participants (n = 15) were randomly selected based on the Trexo size they needed, from a list of those who had previously expressed interest in the study.

### Intervention and Procedure

Participants borrowed a robotic walker for 12-weeks to use in their home or other community settings for at least 5 days/week for 30 minutes/session (150 minutes/week). Caregivers who would be operating the robotic walker throughout the study took part in 3 hours of training over Zoom with staff from Trexo Robotics, where they learned how to use the Trexo, and make any adjustments needed during sessions.

Participants were enrolled for a baseline period of at least 4 weeks before the robotic walking intervention to enable the collection of multiple baseline values (Figure 2) and followed throughout a 12-week intervention and subsequent 12-week follow-up period. Figure 2 represents when each outcome presented in this manuscript was collected.

**Figure. 2:**
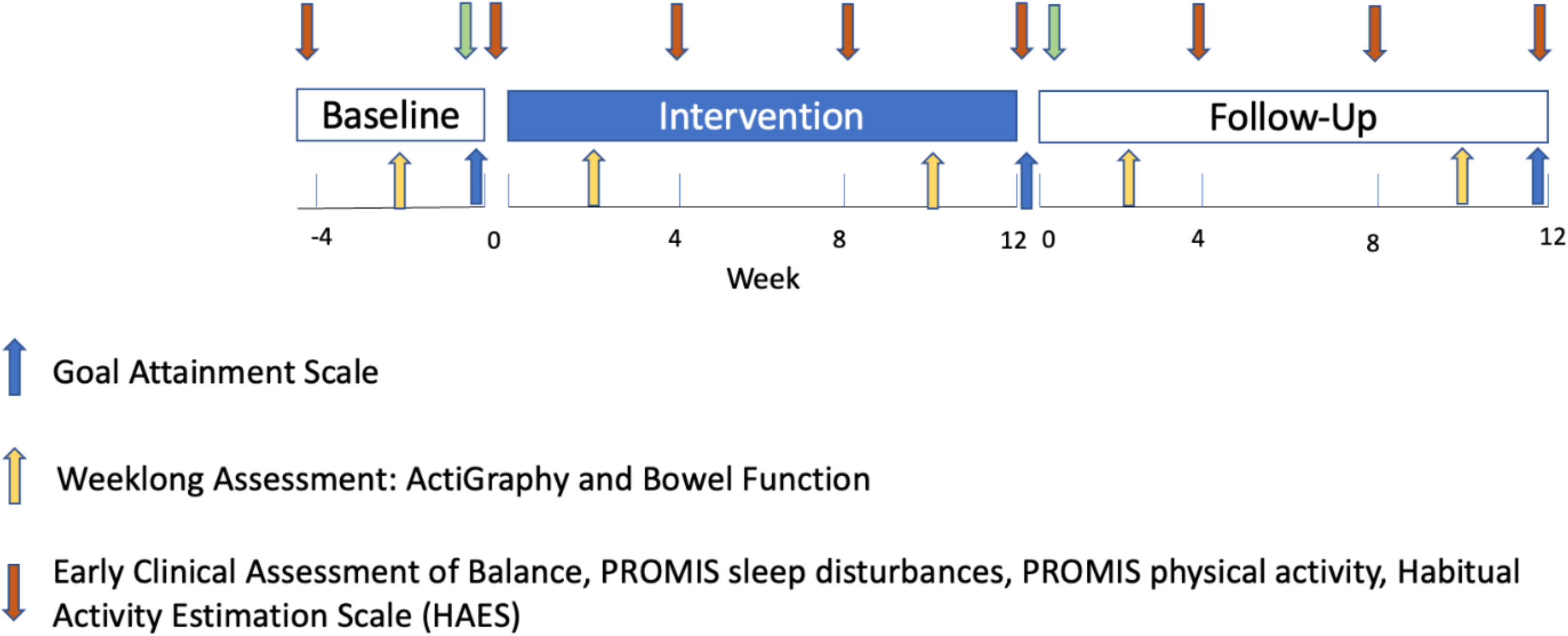
Timeline of interventions showing when each outcome was collected; Abbreviations: PROMIS, patient reported outcome measurement information system

### Quantitative Methodology

#### Outcome Measures

##### Goal Attainment Scale

Clinicians worked with the parents to create measurable goals that they expected to see improvements in following a 12-week robotic walking intervention using the goal attainment scale. Each participant had 1-3 goals depending on how many goals the parents had for their child. These goal assessments were completed before intervention, after intervention, and 12 weeks following intervention. Goals are scored on a five-point scale of -2 to +2 (Cusick et al., 2007). Baseline scores were set at -2 for goals where a decline would not be expected (e.g. physical function) and -1 where a decline could occur (e.g. sleep). The raw scores for every goal for each participant are then transferred into one standardized t-score, where scores of 50 or above indicate that an individual’s goals have been achieved (Bradley et al., 2026; Cusick et al., 2007; Turner-Stokes, 2009).

##### Early Clinical Assessment of Balance

The Early Clinical Assessment of Balance (ECAB) addresses four dimensions of balance across 13 tasks: (1) head and trunk, (2) protective responses for balance in sitting, (3) maintaining upright posture, and (4) making appropriate adjustments for voluntary movements in standing (Westcott McCoy et al., 2019). Each task is rated on a scale of 0-4 to create an overall score. Raw scores are converted into percentiles using developmental trajectories based on the reference population of individuals with cerebral palsy, GMFCS I-IV, and aged 5-12 years old(Chiarello et al., 2021). A clinician within the research group completed the ECAB assessment with each participant before training, every four weeks in the training period and every four weeks in the 12-week follow-up period.

##### Actigraphy Physical Activity Data

ActiGraph wGT3x-BT accelerometer devices (ActiGraph, Pensacola, FL, USA) were used to record the amount (minutes) and percent of time participants spent sedentary, in light physical activity, and in moderate to vigorous physical activity (MVPA) (Hulst et al., 2021). Participants were instructed to wear the accelerometer on their dominant or less impaired wrist for 24 hours per day over a 7-day period(Hulst et al., 2023). Participants were sent home with an activity log to track bouts of activity and times when they took off the ActiGraph (i.e., bathing or swimming). ActiGraphs were initialized to collect data at a resolution of 30Hz. Wear time was validated using the Troiano (2007) equation, where sixty consecutive minutes of zero counts were set as non-wear time (Hulst et al., 2023; Longmuir et al., 2022). Raw accelerometer data were downloaded and converted into 15-second epochs for wrist-worn physical activity and sedentary analysis. Activity intensity was defined by Evanston cut points, as these have been validated in children with cerebral palsy (Clanchy et al., 2011). Participants were asked to wear the Actigraphs prior to training, in the first month of the training period, in the last month of the training period, and after the training period.

##### Habitual Activity Estimation Scale (HAES)

This questionnaire assesses the habitual activity level of children with chronic disorders. It requires parents to recall a typical weekday and a weekend day. Each day is divided into four periods of time (getting out of bed until starting breakfast, breakfast until lunch, lunch until supper, supper until bedtime). Parents fill out the percent of time their child spends being inactive, somewhat inactive, somewhat active, and very active in each of those four periods (Hay & Cairney, 2006). As the original measure was not designed for individuals with severe mobility impairments, we worked with parent-partners and the original creators of this questionnaire to create physical activity definitions that are representative of the lived experiences of the participants in our study. These definitions are as follows: Inactive: any activity done without moving and fully supported (e.g., napping, reading, watching television, lying down), Somewhat inactive: any activity done without moving but supporting some of your body (e.g., watching television sitting up, reading, video games, sitting in school, commuting to/from school), Somewhat active: activity done which involves movement and increases heart rate or makes it a little hard to talk (e.g., walking with an assistive device, playing catch, helping with chores, standing frame), Very active: any activity which causes one to sweat, heart rate to increase dramatically, causes heavy breathing and makes it very difficult to talk (e.g., playing sledge hockey or other sports, swimming, dance). Parents filled out this questionnaire before the intervention started, and then every four weeks after that until the 12-week follow-up period.

##### Patient Reported Outcome Measurement Information System (PROMIS)

The Patient Reported Outcome Measurement Information System (PROMIS) is a set of standardized questionnaires that measure how individuals feel in key areas of their health (Hooke et al., 2021). In this study, we used the parent-reported versions of the PROMIS Physical Activity Short Form 4a and PROMIS Sleep Disturbance Short Form 8a Questionnaires. Raw scores from these questionnaires are converted into a standardized T-Score metric, such that the mean of the reference population is 50 (Hooke et al., 2021). Parents filled out this questionnaire before, every four weeks during the training period and every four weeks in the 12-week follow-up period. The PROMIS Physical Activity Questionnaire examines a 7-day reporting period (Hooke et al., 2021). In this questionnaire, a higher score means the participant is getting more physical activity. The PROMIS Sleep Disturbance questionnaire is designed to reflect the lived experiences of individuals experiencing sleep difficulties (Forrest et al., 2018). A higher score reflects higher sleep disturbances and indicates poor sleep health. Severity of sleep disturbances are categorized as t-scores of: <55 = normal limits, 56-59 = mild, 60-65 = moderate, >66 = severe (Forrest et al., 2018).

##### Bowel Function Diaries

Participants were given a week-long bowel function diary developed with our patient partners before intervention, during the first month of the training period, during the last month of the training period, and during the first month following training. For each bowel movement during the week, parents were asked to report when their child had a bowel movement and the Bristol Stool Scale. Interventions to support bowel function (e.g. medication, dietary strategies or procedures such as an enema) were tracked separately. Participants were classified as constipated or not at each time point. Constipation was defined as: 2 or fewer bowel movements a week, Bristol Stool Scale 1 or 2 for the majority of bowel movements that week, or taking medications to facilitate bowel movements.(Cankurtaran et al., 2023; Lauruschkus et al., 2022) At each time point following the baseline measure, bowel function was classified as improved, no change, worse or cannot be determined compared to the participant’s baseline bowel function diary (Figure 3).

**Figure 3:**
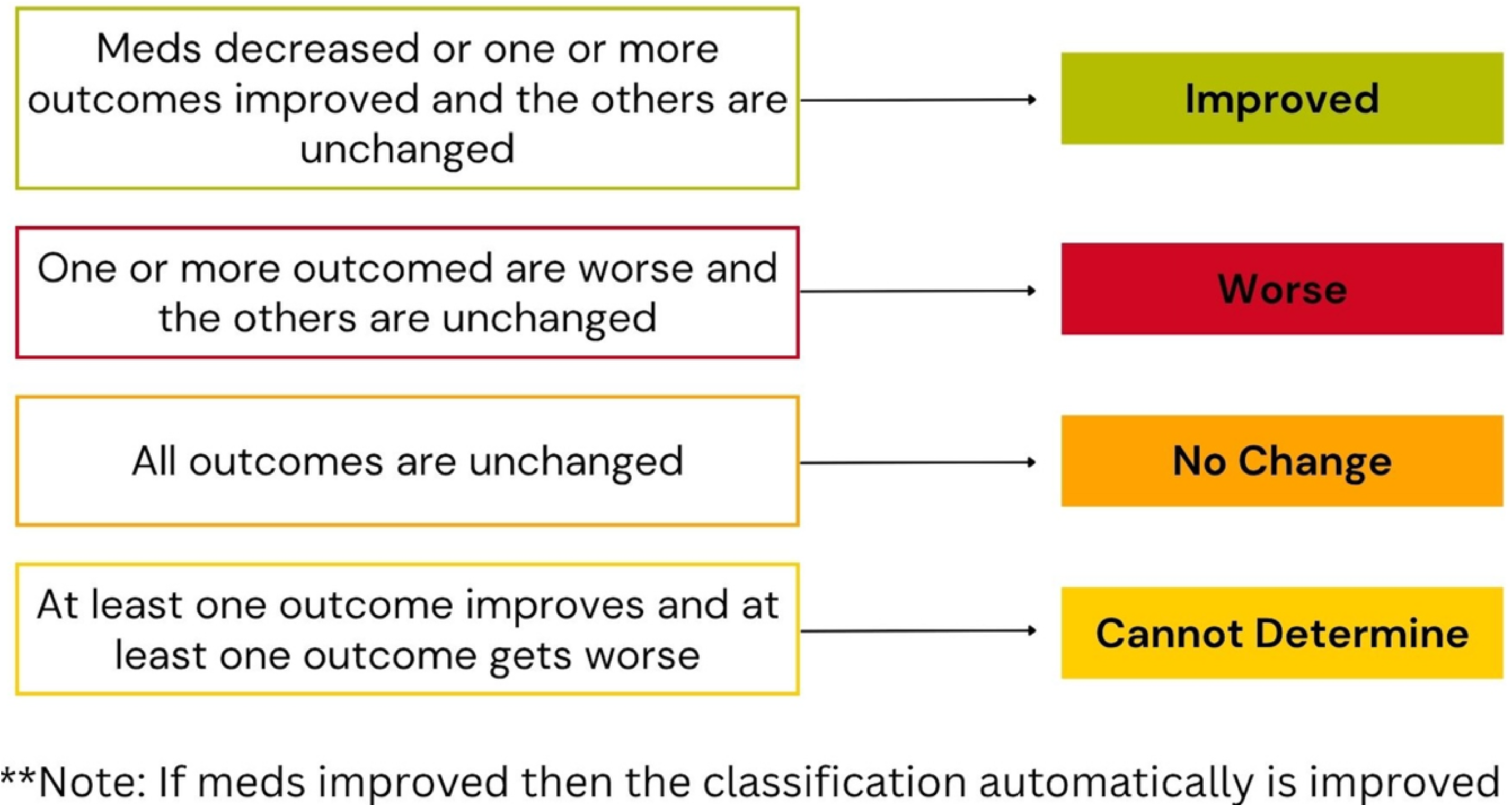
How decisions regarding if bowel function improved or not at each time point were made

#### Quantitative Analysis

Given our small sample size, we conducted non-parametric statistics (median (25th – 75th percentiles)) for all outcome measures. RStudio (Posit Software, Boston, Massachusetts v.2026.01.0+392) was used to conduct a Skillings-Mack test of difference to assess the difference in scores across each time point. Following a significant Skillings-Mack test, a post-hoc Wilcoxon Signed-rank test was used to determine which time points had a significant change. An exploratory Wilcoxon signed-rank test was conducted on outcomes where there was a non-significant Skillings-Mack test, but visually, it appeared certain time points may be significant. The Skillings-Mack test was used because it does not exclude participants who are missing data at certain time points. Time points with missing data were left blank in the analysis as the sample size was too small for other techniques used to address missing data. An association test (Spearman’s correlation) was used to examine associations between outcomes and the participants’ usage and demographic details. An alpha of 0.05 was used for all outcome measures.

### 3.3.6 Qualitative Methodology

Qualitative data were collected in this study in order to understand if the quantitative findings varied from what parents were noticing and to understand if there were any areas where parents saw improvements that we did not measure.

#### Theoretical Framework

The qualitative portion of this study was guided by a relativist ontology and constructivist epistemology. A relativist ontology acknowledges that each individual has their own reality, which is constructed based on their lived experiences (Dieronitou, 2014). A constructivist epistemology acknowledges that knowledge is created through interactions with others and understanding the lived experiences of others. Therefore, the interviewer interacting with the participants and understanding those lived experiences creates new knowledge. This philosophical approach allowed the researchers to build knowledge regarding the commonalities and differences in the experiences of families as they relate to using the robotic walker. Further, this approach recognizes that each family will have had different experiences and interactions, all of which will be reflected in their “truth” (Dieronitou, 2014).

#### Methodology

Qualitative description was the overarching methodology used in this study. One of the main tenets of this approach is to stay close to the data to create a rich description of how participants experienced an event, making it ideal for mixed-method studies. Like most qualitative methodologies, this approach is inductive in nature to describe participants’ experiences that address the research question (Neergaard et al., 2009; Sandelowski, 2000).

#### Semi-Structured Interviews

Semi-structured interviews were conducted before and after the intervention period to understand areas of physical function where parents were hoping to see improvements and to understand if parents saw improvements in these and other areas of physical function and how these improvements impacted their lives on a daily basis. Interviews were conducted via videoconference (Zoom 6.7.7; Zoom Communication; San Jose, CA, USA)) with parents or guardians, and participants if they were willing and capable. The interview guide asked about areas where parents saw improvements and new things their child does now that they may not have done before, and how these changes impacted activities of daily living (Appendix A).

#### Qualitative Analysis

Interviews were audio-recorded and transcribed verbatim by Zoom. All transcripts were read through thoroughly and anonymized before analysis began. NVivo12 software (QRS International Pty Ltd.) was used to store and analyze data. Data were analyzed using thematic analysis. The thematic analysis was conducted in six steps: (1) Transcripts were read and re-read to gain familiarity with the data, (2) Re-reading of transcripts to generate initial codes, (3) Codes were grouped to create themes across the dataset, (4) Themes were reviewed and checked to ensure they aligned with the lived experiences of the participants, (5) Names and definitions of codes were created, (6) Finally, the report was written (Braun & Clarke, 2019, 2020). This analysis method was useful for this study as it allowed the researcher to further understand and compare the lived experiences of the participants.

## Results

### Demographics

There were 15 participants (8 males, 7 females) who completed this study. The median age of participants was 6.5 (5.3 -10) years, as previously reported in a related paper describing the intervention feasibility and impacts on families (Youngblood et al., 2026). The majority of participants had cerebral palsy (10/15) and were able to take some steps with partial weight bearing (Gillette Functional Assessment Questionnaire (FAQ) 2) and used a wheelchair to ambulate in community settings (GMFCS IV) (Table 1). All participants had a bilateral distribution of their motor impairment. They also had a variety of motor impairments, including mixed spastic-dyskinetic dystonic (7/15), athetosis (1/15), spastic (4/15) and hypotonic (3/15). Participants also had a variety of aetiologies of their diagnoses, including hypoxic-ischaemic encephalopathy (5/15), infectious (2/15), periventricular white matter injury associated with prematurity (2/15), multifocal strokes (2/15) and genetic factors (5/15). Note, one participant had multiple aetiologies.

**Table 1:**
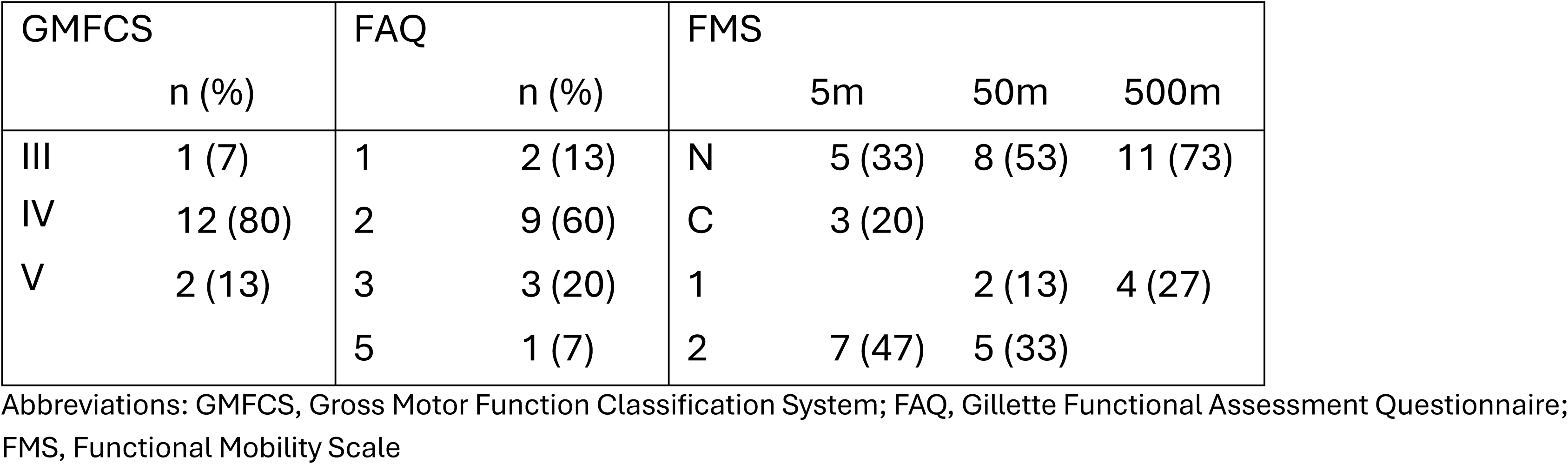
Participant ambulatory ability information.

### Feasibility of Study Outcome Measures

All participants scheduled the mandatory in-person assessments (GAS and ECAB), but changes in health status (e.g. surgery and broken bone) resulted in missing data points for these outcomes (n = 2). Families could opt out of the other outcomes in an attempt to reduce the burden on families. Figure 4 shows the number of participants who completed each outcome at all the timepoints at which they were collected. Adherence to Actigraphy use presented challenges due to participant discomfort and device-related difficulties. There were also difficulties with bowel movement diary adherence, as parents reported difficulties with tracking their child’s bowel movements for a whole week. Parents also reported study fatigue, resulting in missing data during the follow-up.

**Figure 4:**
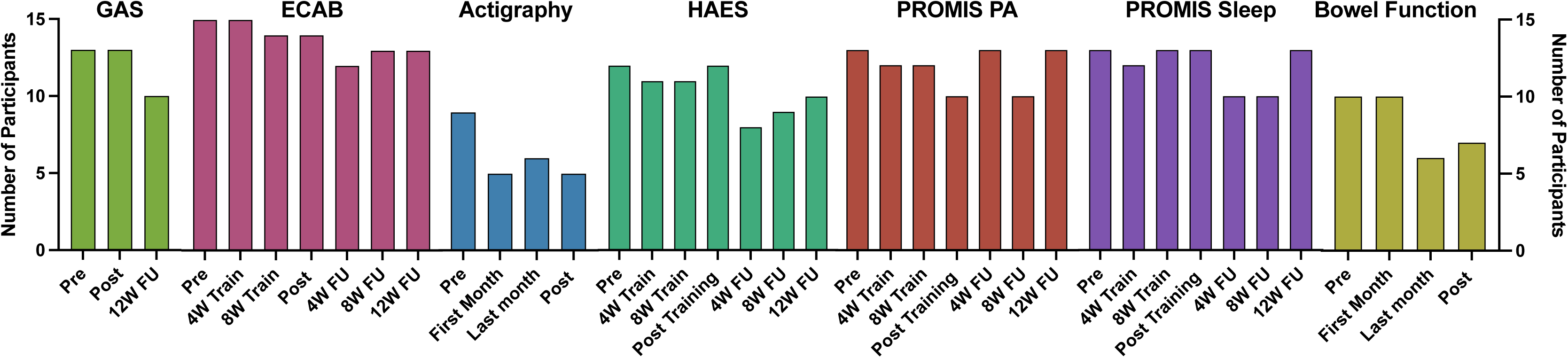
The number of participants who filled out each outcome at all timepoints they were collected. Abbreviations: GAS, goal attainment scale; ECAB, early clinical assessment of balance; HAES, habitual activity estimation scale; PROMIS, patient reported outcome measurement information system

### Goal Attainment Scale (GAS)

Most participants had goals related to functional mobility (n = 8) and personal care (n = 7). Other categories of goals were socialization (n = 4), active recreation (n = 3), school and/or play (n = 2) and quiet recreation (n = 1). 10/13 participants achieved their goals (T-score ≥ 50) after the intervention period, and 5/10 maintained their goal achievement at the follow-up period (Figure 5).

**Figure 5:**
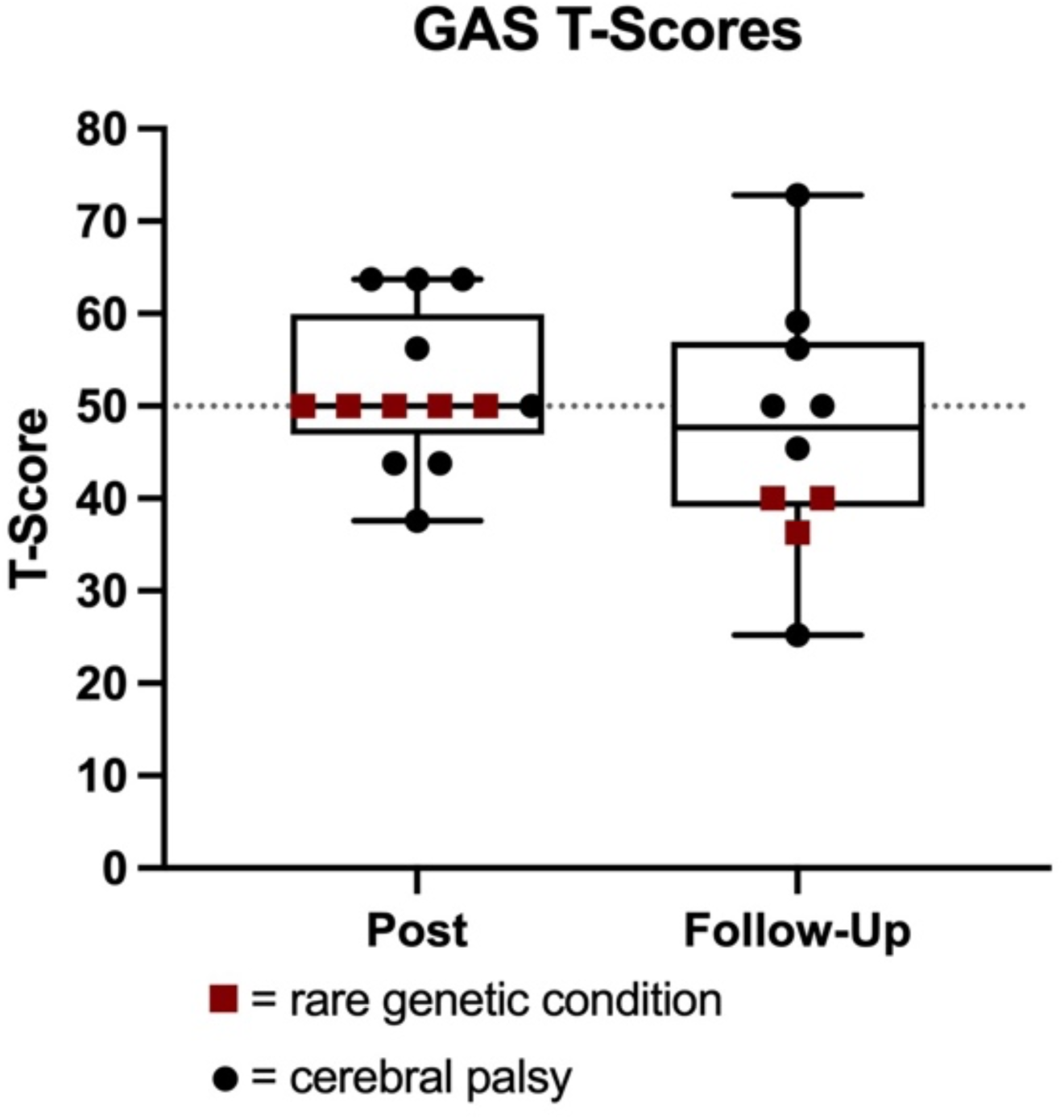
Participants’ T-Scores post-training and 12-week follow-up periods. A score of 50 or above means the participant’s goals were achieved. Abbreviations: GAS, goal attainment scale

### Early Clinical Assessment of Balance

There was a significant improvement in the ECAB across time points (p = <0.001, Skillings-Mack Statistic = 23.08) (Figure 6). These improvements became significant after 8-weeks of training (p = 0.016, change in percentiles = 5.0 (0.0-21.4)), and were sustained through to the 12-week follow-up time point (p = 0.008, change in percentiles = 13.7 (3.1 – 23.7). Participants who walked for longer durations (minutes) (p = 0.039, R^2^ = 0.310) and took more steps (p = 0.046, R^2^ = 0.292) experienced a significantly larger change in ECAB at the post-training time point (Appendix B; Supplement Figures 1 & 2). These correlations were no longer significant at the 12-week follow-up time point (minutes: p = 0.579, R^2^ = 0.028; steps: p = 0.494, R^2^ = 0.043). Participants with greater ambulatory abilities (assessed using FAQ) had larger changes in post-training ECAB scores (p = 0.005, R^2^ = 0.529), and this correlation was no longer significant at the 12-week follow-up time point (p = 0.823, R^2^ = 0.005) (Appendix B; Supplement figures 1 &2).

**Figure 6:**
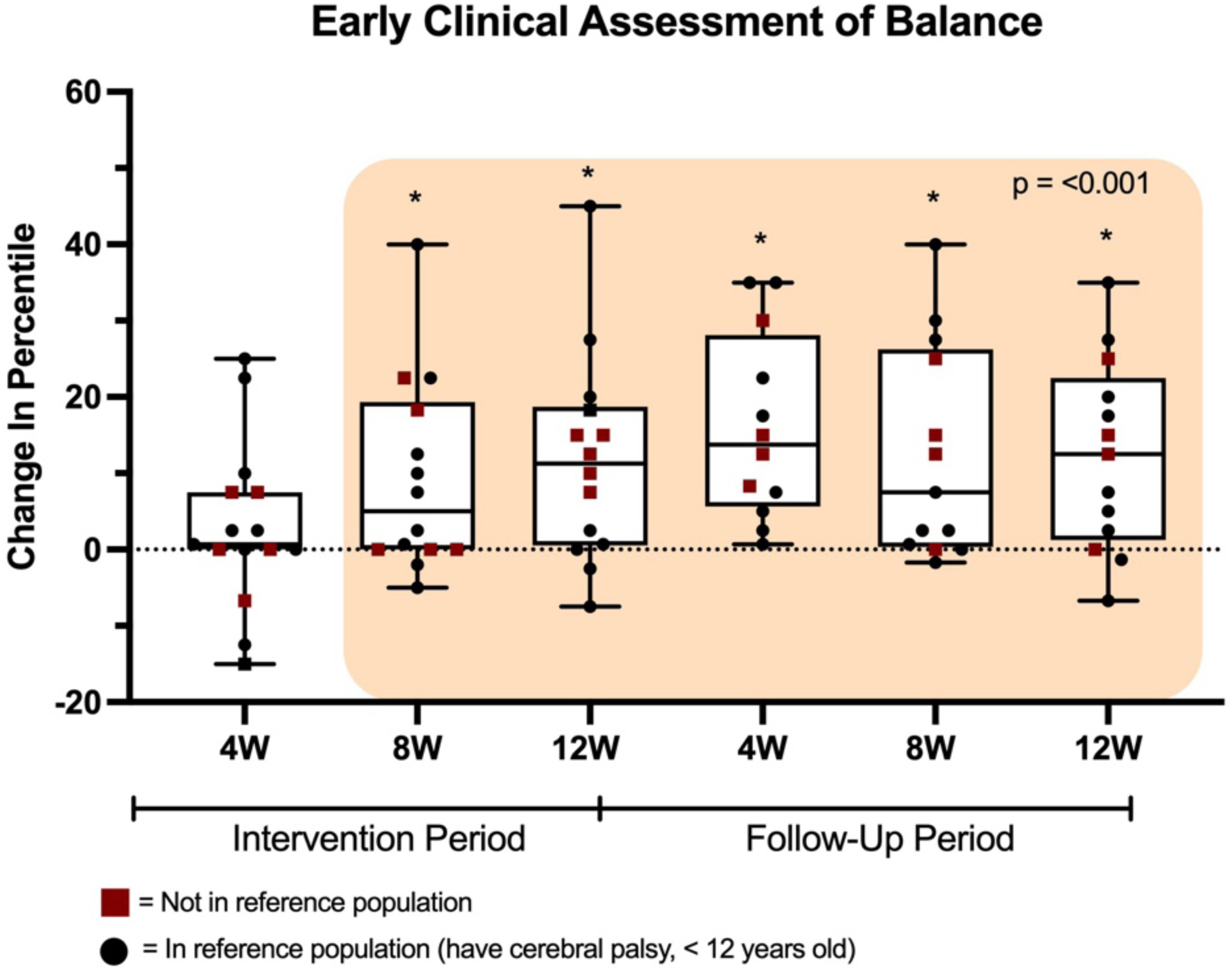
Changes in ECAB scores across the intervention and follow-up periods. Boxplots in orange square mean change was significant at that time point

### Actigraphy Data

There were no significant changes in the percent of time spent sedentary, at light physical activity, or at moderate to vigorous physical activity (MVPA) at any time points (Table 2). There was also no significant change in average daily minutes spent sedentary, at light physical activity, or at MVPA at any time points throughout the intervention and post-training period (Table 3). Wear time (in minutes/day) at each time point is as follows: pre - 768 (574 – 1025), training period - 780 (691 – 848), post – 809 (674 – 998). After cross-referencing the actigraphy results and tracking of device usage, we found that robotic walking was registering as sedentary time on the ActiGraphs, likely because they were worn on the wrist.

**Table 2:**
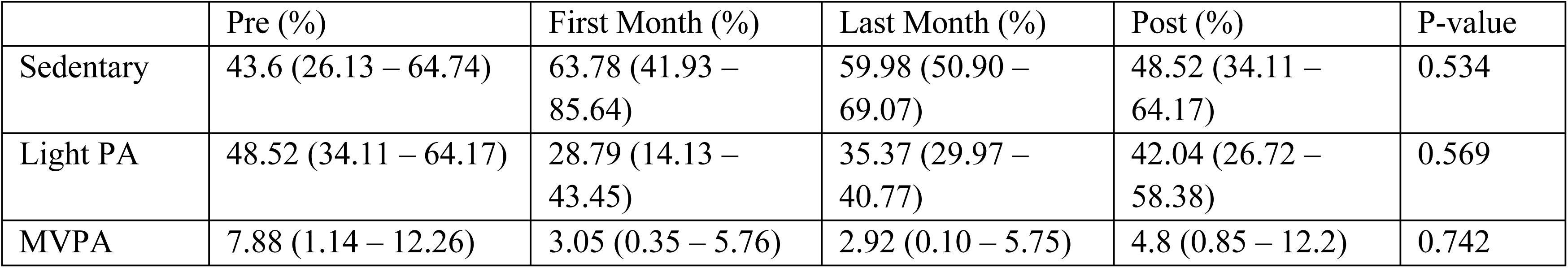
Percent of time participants spent sedentary, in light physical activity (PA), and in moderate to vigorous physical activity (MVPA). Descriptive statistics are represented as median (25^th^ – 75^th^ percentile).

|  | Pre (%) | First Month (%) | Last Month (%) | Post (%) | P-value |
| --- | --- | --- | --- | --- | --- |
| Sedentary | 43.6 (26.13 – 64.74) | 63.78 (41.93 – 85.64) | 59.98 (50.90 – 69.07) | 48.52 (34.11 – 64.17) | 0.534 |
| Light PA | 48.52 (34.11 – 64.17) | 28.79 (14.13 – 43.45) | 35.37 (29.97 – 40.77) | 42.04 (26.72 – 58.38) | 0.569 |
| MVPA | 7.88 (1.14 – 12.26) | 3.05 (0.35 – 5.76) | 2.92 (0.10 – 5.75) | 4.8 (0.85 – 12.2) | 0.742 |

**Table 3:** Daily average of minutes spent sedentary, at light physical activity (PA), and at moderate to vigorous physical activity (MVPA). Descriptive statistics are represented as median (25th - 75th percentile).

|  | Pre (min/day) | First Month (min/day) | Last Month (min/day) | Post (min/day) | P-value |
| --- | --- | --- | --- | --- | --- |
| Sedentary | 304 (204 – 513) | 406 (280 – 532) | 278 (117 – 440) | 489 (164 – 543) | 0.144 |
| Light PA | 304 (247 – 613) | 567 (405 – 729) | 342 (307 – 377) | 260 (241 – 358) | 0.309 |
| MVPA | 55 (9 – 82) | 30 (3 – 57) | 25 (0.4 – 49) | 57 (7 – 82) | 0.526 |

### Habitual Activity Estimation Scale

There was no significant change in parent-reported percent of time spent active on weekdays (p = 0.436, Skillings-Mack Statistic = 5.88) or weekends (p = 0.177, Skillings-Mack Statistic = 8.92) throughout the intervention and follow-up periods (Appendix B; Supplement Figure 3).

### Patient Reported Outcome Information System (PROMIS) Physical Activity

There was no significant change in parent-reported physical activity throughout the intervention and follow-up (p = 0.128, Skillings-Mack Statistic = 9.92) (Appendix B; Supplement Figure 4).

### Patient Reported Outcome Information System (PROMIS) Sleep Disturbances

Prior to the intervention, over half of the group (7/13) experienced severe sleep disturbances, which decreased to less than one-third (4/13) post-training (Figure 7). There were no significant changes in the PROMIS sleep disturbance questionnaire across all time points (p = 0.343, Skillings-Mack Statistic = 6.76). However, there was a clear U-shaped pattern showing potential localized benefit to sleep at the post-training and 4-week follow-up time points, which is no longer present as the follow-up period continues (Figure 8). Given the exploratory nature of the study and the risk of falsely accepting the null hypothesis with our small sample using a Skilling-Mack Test, we chose to run additional Wilcoxon signed rank tests to assess the data through all possible avenues. This revealed significant changes directly after the intervention period (post-training (12W) (p = 0.025, change = -2.3 (-8.55 - -1.35)) and at the 4-week follow-up time point (p = 0.047, change = -3.25 (-4.3 - -1.7)).

**Figure 7:**
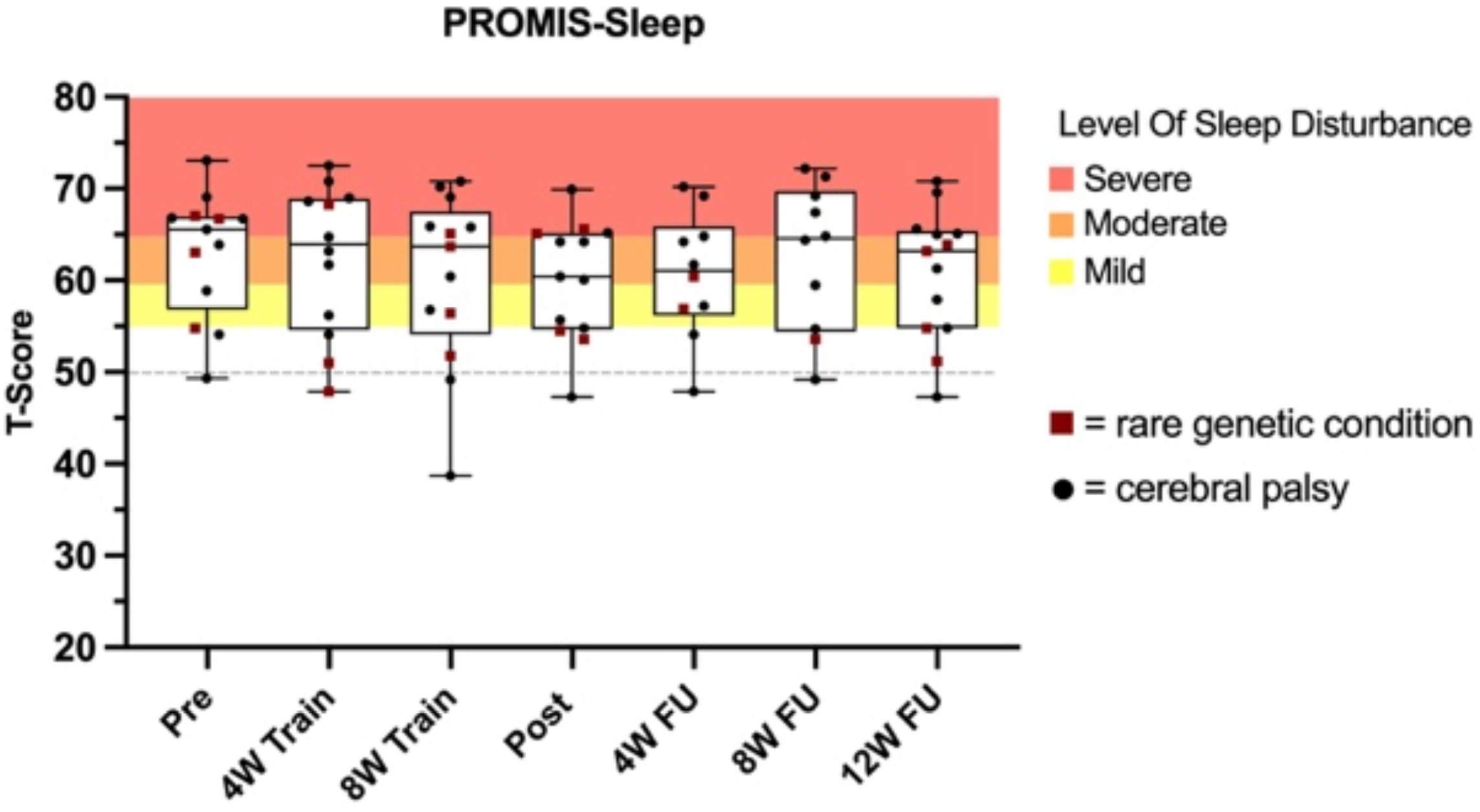
PROMIS Sleep disturbance scores at each time point. A score of 50 is the average score of the population

**Figure 8:**
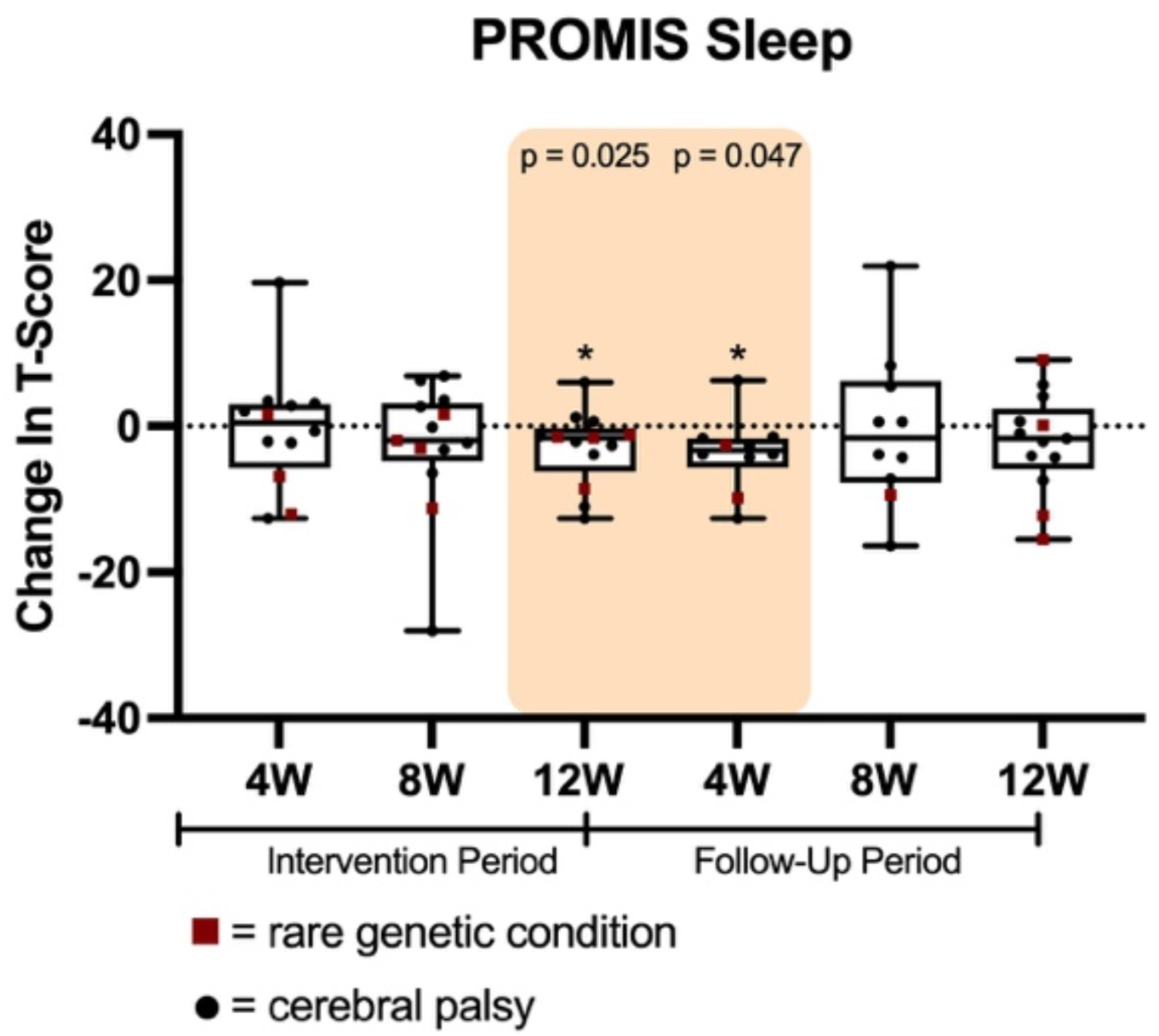
Change scores in the PROMIS sleep disturbances questionnaire at each time point. Time points in the orange box = improved

### Bowel Function

There were 10/15 participants who completed weeklong bowel movement diaries at baseline. Out of these participants, 7/10 were constipated at baseline. All participants who were not constipated at baseline remained not constipated throughout the study. Four of the 7 participants who were constipated at baseline had improvements in bowel function in the first month of training. Only 5 were reported in the last month, and a different 5 were reported post-training; 3/5 improved after the intervention period (all three participants were no longer constipated after the training period). While no one got worse, not enough participants reported data at the subsequent time points to draw any general conclusions. The results of all the constipated participants are reported in Table 4.

**Table 4:** Changes from baseline in bowel function in participants who were constipated at baseline. * = participant is not constipated at that time point. Black spaces mean missing data. Note – participants could still be classified as improved in any metric (frequency, Bristol, or medication), but they may still be constipated for other reasons.

| Participants | First Month of Intervention | Last Month of Intervention | Post Intervention |
| --- | --- | --- | --- |
| 01 | Cannot Determine | Improved | Improved* |
| 04 | Improved |  | Improved* |
| 03 | Cannot Determine | No Change | No change |
| 06 | Worse | No Change | No change |
| 11 | Improved | Improved | Improved* |
| 15 | Improved* |  |  |
| 18 | Improved | No Change |  |

### Qualitative Results

Nine families completed pre- and post-training interviews. This included seven families in which one parent was interviewed, one family in which two parents were interviewed together, and one in which the participant and a parent were interviewed together. Three themes were identified where families noticed improvements in: 1) standing and walking, 2) activities of daily living, which they frequently attributed to increased strength and/or increased head and trunk control, and 3) sequelae of inactivity (Figure 9).

**Figure 9:**
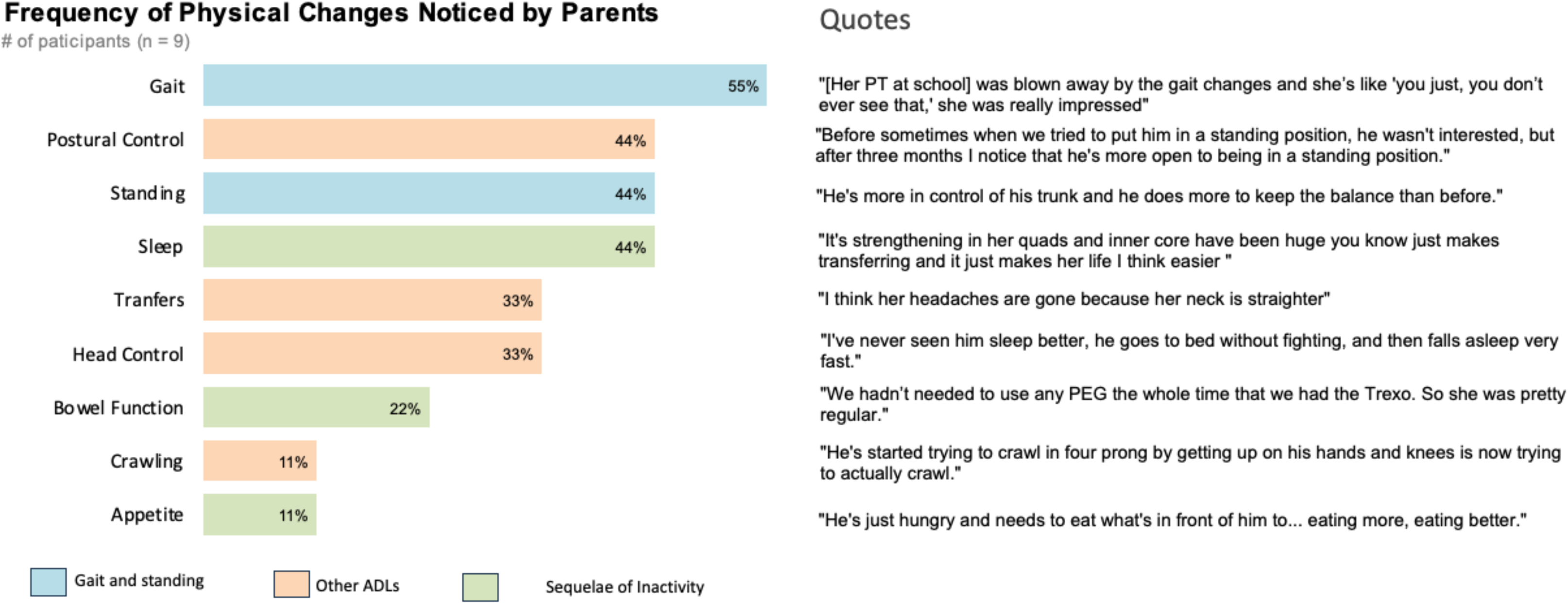
How many parents mentioned certain topics and quotes to support these results

### Theme #1 – Families noticed an improvement in the walking pattern, willingness to walk, and stand

Many parents reported seeing an improvement in their child’s gait pattern and noted that their child was more willing to use their walker and stand for longer after the intervention. When discussing this improvement, one father mentioned, “We did notice that his movement patterns were better, so his legs were tracking with his feet not turned in and actually in line, and his knees tracking properly. And we’ve noticed a bit of regression already since not using it, with his knees collapsing in again and his feet turning in” (TR18 Dad). Families feel this resulted in safer walking with less risk of falling. Parents also mentioned how practitioners have commented on how much their child’s gait has improved, with one mom stating, “Her school PT who was there initially with helping… she had seen [TR04 name] walking before and then she wasn’t really around much during the whole time with the Trexo and then she saw her again recently in her walker, and she was blown away by the gait changes and she’s like ‘you just, you don’t ever see that,’ she was really impressed” (TR04 Mom).

In the pre-training interviews, many parents mentioned how they were excited for their child to walk more, with one mom stating, “I couldn’t imagine what it would be like to spend a whole day sitting. You know that as soon as people are done surgery in a hospital, the first priority from a health standpoint is to get up and get moving, get ambulatory, so to give her that opportunity, I think, has huge impacts on her life” (TR04 Mom). After the intervention, many parents reported that their child’s endurance and willingness to walk had increased greatly, which has led to more walking. When discussing how their child enjoys walking more now, one mom mentioned, “He wants to try to walk more to, even if it’s just with us, he wants to walk more” (TR03 Mom). Additionally, the same parent mentioned how their child has made gains in other mobility, “He’s started trying to crawl in four prongs by getting up on his hands and knees is now trying to actually crawl” (TR03 Mom).

Parents believe that many physical changes in their child have led them to enjoy standing and walking more. These changes include increased trunk control, stronger head and neck control, and greater leg strength. In pre-training interviews, most parents mentioned that their children did not enjoy using their standing frame before the intervention, and this caused stress on the parents. After the intervention, one mom mentioned, “She likes her standing frame a lot now, whereas before she didn’t, so she can stand easily 2 hours in her standing frame, whereas she couldn’t before. And that might just be because she has more trunk control and it’s less exhausting on her neck, things are strong, everything’s stronger” (TR04 Mom). Parents also mention how their child will now stand when watching TV instead of sitting. Their child standing more makes parents feel better and less worried because they know their child is experiencing “better circulation,” “an increase in strength and muscle mass” and “bone density” benefits while standing. When discussing the benefits of standing, one dad mentioned, “…it’s good because we know that [standing] brings benefits for his hips” (TR05 Dad). One mom also mentioned how her daughter, “…seems healthy and her circulation is better standing in her Walker” (TR01 Mom). Overall, parents have noticed an improvement in their child’s gait, and their child is more willing to walk and stand more. This has led parents to feel less worried about getting their child to walk or stand enough for therapeutic benefits.

### Theme #2 – Children showed improvements that impacted a variety of activities of daily living

Many parents noticed an increase in overall strength. that impacted how the children were able to participate in activities of daily living. Managing aspects of their child’s lives became easier because the children were able to help more. When discussing the increase in strength, one mom mentioned, “It’s so cool how much strength she gained in those three months, it was shocking that you’re like ‘oh man, the sky is the limit’ if you keep using this” (TR04 Mom). Parents have also noticed their child has an increase in muscle mass now, with one mom stating, “She looked more bird-like previously, and she looks more kid-like now, for lack of a better word, her muscles look appropriate to her bones, whereas before they were more atrophied looking” (TR04 Mom).

Many parents noticed these increases in strength frequently resulted in easier transfers. Improvements in transferring were also important to parents in the pre-training interviews. One dad discussed how he was hoping to see his child gain more independence with transfers by stating, “…any benefit to independence is going to take strain off ourselves, it takes strain off her caregivers, people at school, anyone who helps with her daily life, so really that’s anything that can take the physical strain off us and more importantly give her independence” (TR06 Dad).

When discussing how transferring her child is easier following training, one mom mentioned, “And even for everyday things like transferring, her legs are so much stronger she now instead of having to pull her up…she just pops up” (TR01 Mom). One parent also mentioned how their child is getting out of bed on their own now. They stated, “He’s been getting out of bed himself a lot more lately… And when he was little, he used to get out of bed a lot, but I think as he got bigger, he was tighter it felt a little scarier to get out of bed, but almost every morning, when I go in to wake him up, he’s already on the floor playing. So that’s new. I don’t know if it’s related, but it’s new” (TR11 Mom).

Parents also noticed an increase in head and trunk control, which has helped with certain aspects of their daily life. When discussing how her child now has more head control, one mom mentioned, “…she spends less time slumped over and can make eye contact with people, and smile at more people.” (TR04 Mom). When discussing how much easier it is to get his child dressed now due to an increase in trunk control, one dad mentioned, “Yeah, for example, when I normally sit him on the bed when I’m dressing him up…when I put the T-shirt on, he normally goes to the side. I always hold him, like his legs, but now when he goes to the side, he can go back to centre on his own” (TR14 Dad).

### Theme #3 – Parents noticed improvements in areas related to a sequelae of inactivity

Parents noticed improvements in body functions associated with sedentary lifestyles, such as bowel function, appetite and sleep. In the pre-training interviews, many parents mentioned how they were hoping to see an improvement in their child’s bowel movements or sleep. In the post-training interviews, these parents were excited that they saw an improvement in bowel function during the intervention. When discussing how they were able to decrease medications to facilitate bowel movements, one mom stated, “We hadn’t needed to use any PEG (Polyethylene Glycol – medication typically used for constipation) the whole time that we had the Trexo, PEG being a stool softener. So, she was pretty regular. She often liked to poop while she was in the Trexo” (TR04 Mom). One dad also mentioned how he was happy that his child is eating better now, by stating, “…he goes out and does one of his Trexo walks comes home. He’s hungry, and you know again he’s eating everything in front of him, being less picky because he’s just hungry and needs to eat what’s in front of him” (TR14 Dad). Children who previously struggled with sleep slept better after using the intervention. When discussing what this improvement in sleep means to him, one dad mentioned, “I mean, when he’s falling asleep quickly and getting up and not fighting. Obviously, he’s just getting better sleep. And you know, as any parent will tell you, when your 6-year-old’s not getting sleep. Your life is not fun” (TR14 Dad).

## Discussion

The key finding of this study indicates that postural control improved after 8-weeks of a 12-week robotic walking intervention, and this improvement was sustained through the 12-week follow-up period. Other measured outcomes that improved immediately following the intervention were not sustained. Specifically, the majority of participants reached their goals after the intervention period, but these results were not sustained at the 12-week follow-up assessment. It’s important to note that while GAS goals did not have an overall longer-term improvement, only 1 participant had a t-score below their baseline, and the rest were either at or above their baseline scores at the 12-week follow-up period. Sleep also improved after 12-weeks of robotic walking but was only maintained 4-weeks into the follow-up period. There were no clear improvements in objective measures of physical activity and bowel function. Parents mentioned improvements in other areas, as well as improvements in strength and walking ability.

Participants’ improvements in postural control were consistent with the improvements parents reported in activities of daily living. Similar improvements in postural control have been shown in a variety of robotic walking studies in other populations (Khan et al., 2019; Tsai et al., 2021) and in children with cerebral palsy, as summarized in a recent meta-analysis (Chen et al., 2026; Wang et al., 2024). In RoWaCaWa, we found significant improvements in the ECAB, particularly in areas related to seated balance. Parents discussed improvements in trunk and head control that improved aspects of their daily routine, such as transfers and dressing. Further, many participants also improved in predefined goals related to postural control. This may decrease caregiver strain as non-ambulatory children require more hands-on support from their parents for transfers, positioning, and other activities of daily living (Noritz et al., 2022).

Participants in this study had subjective improvements in outcomes related to walking and standing. This is consistent with objective systematic reviews involving multiple devices and populations (Lee & Kim, 2025; Postol et al., 2026). However, these studies mainly examine gait impacts on children GMFCS levels II – III, with a few individuals GMFCS level IV included in the studies (Postol et al., 2026). While objective measures of gait are challenging to collect in individuals with severe mobility impairments (e.g. GMFCS levels III-V), the subjective measures used in this study are particularly relevant, as they were shown through improvements in parents’ predefined goals and their additional observations. These improvements in walking may improve the quality of life in both children with mobility impairments and their caregivers, though these benefits were not seen using the assessment tools we chose, as previously reported (Mutoh et al., 2019; Youngblood et al., 2026). The results in our study present the importance of including children with severe mobility impairments in robotic walking interventions and selecting assessments of walking, stepping and standing to further understand how these interventions can improve walking abilities.

There were no measured changes in objective measures of physical activity. When examining the actigraphy data, physical activity (both light and MVPA) appears to decrease while sedentary time appears to increase. This may have occurred because families were replacing other activities with robotic walking. Future studies that want to use actigraphy to examine physical activity from a robotic walking intervention may benefit from using hip-worn actigraphy while the participants are in the robotic walker (Longmuir et al., 2022). While the objective measures of physical activity did not show improvements, in the qualitative interviews parents did mention that their child is standing and walking more now, but these changes were not picked up from the objective measures we used.

In regard to sequelae of inactivity, the results from objective measures were unclear. Missing data confounded the interpretation of the bowel function data. An exploratory analysis revealed an improvement in sleep at the post-training and 4-week follow-up time points, and these results are similar to a large observational study in which improvements in sleep were observed (Hilderley et al., 2026). Bowel function has not been studied widely in the current robotic walking literature, and more studies are needed to understand the exact impact. While the objective measures did not show clear results parents did mention improvements in sleep and bowel function.

Overall, robotic walking showed clear improvements in postural control (measured by ECAB), measured goals (measured by GAS) and sleep disturbances. It is important to note that improvements in postural control were sustained throughout the 12-week follow-up period, whereas improvements in goals and sleep disturbances were not. This is typical, as the benefits of a walking/physical activity intervention would not be expected to be sustained once walking stops (Liang et al., 2026; Wolan-Nieroda et al., 2022). Parents discussed how their child stands and sits unsupported more now; these improvements may be the reason why improvements in postural control were sustained through the follow-up period when other outcomes were not. As balance is improved the most when it is incorporated in activities of daily living (Noritz et al., 2022), this may explain the persistence of benefits observed in this study.

## Limitations

As this is an exploratory single-arm study, it generates a low level of evidence. Since the majority of our participants had cerebral palsy and were GMFCS level IV, generalizability might be limited to that or similar populations, and these results may not represent the lived experiences of all individuals who use a robotic walker. We may have underestimated the changes in GAS goals, as we had challenges setting an expected outcome, as outcomes were previously unknown. The use of outcome measures that are not specifically validated in this population may have impacted our ability to detect impacts, as was highlighted by parents who found that some questions did not reflect the lived experiences of their children. Different measures of physical activity are needed to provide a more accurate measure of how robotic walking impacts physical activity. Missing data were prevalent throughout the study. Missing data occurred for several reasons: changes in participant health status, misplaced assessments (bowel movement diaries and questionnaire), and parents experiencing study burnout and were therefore unable to complete all outcomes. Since the impacts of robotic walking are wide-ranging, we attempted to investigate many of these impacts, but as a result, this impaired our ability to collect all outcome measures due to study burnout. The Skilling Mack test did not detect an overall significant improvement in sleep disturbances, and the pairwise findings (Wilcoxon Signed Rank) should be considered exploratory and hypothesis-generating.

## Conclusions

The results of this study suggest that robotic walking is an effective rehabilitation intervention for improving postural control and family goals in ways that are relevant to daily life. Parents also reported improvements in strength and balance that enhanced activities of daily living and positive changes to the bowel and sleep impacts of sedentary lifestyles. To our knowledge, this interventional study is the largest examination of the physical function impacts after using a robotic walker in a home and community environment. However, controlled studies are needed to increase the level of evidence. This work can be used by parents and clinicians who are interested in using robotic walking to improve aspects of physical function in their child or patient.

## Data Availability Statement

Most of the data that support the findings of this study are available openly; some data is held under restriction to protect participant privacy. Access to the open data and instructions for accessing the data are available on the “RObotic WAlking for children who CAnnot WAlk (RoWaCaWa): Impacts on Physical Function and Physical Activity from a 12-week robotic walking intervention”, https://doi.org/10.5683/SP4/JHV2OK, Borealis, DRAFT VERSION.

